# Number of COVID-19 hospitalisations averted by vaccination: Estimates for the Netherlands, January 6, 2021 through August 30, 2022

**DOI:** 10.1101/2022.12.20.22283713

**Authors:** Senna C.J.L. van Iersel, Scott A. McDonald, Brechje de Gier, Mirjam J. Knol, Hester E. de Melker, C.H. (Henri) van Werkhoven, the RIVM COVID-19 epidemiology and surveillance team, Susan J.M. Hahné

## Abstract

**Background:** Vaccines against COVID-19 have proven effective in preventing COVID-19 hospitalisation. In this study, we aimed to quantify one aspect of the public health impact of COVID-19 vaccination by estimating the number of averted hospitalisations. We present results from the beginning of the vaccination campaign (period 1, January 6, 2021) and a period starting at August 2, 2021 (period 2) when all adults had the opportunity to complete their primary series, until August 30, 2022.

**Methods:** Using calendar-time specific vaccine effectiveness (VE) estimates and vaccine coverage (VC) by round (primary series, first booster and second booster) and the observed number of COVID-19 associated hospitalisations, we estimated the number of averted hospitalisations per age group for the two study periods. From January 25, 2022, when the indication of hospitalisation was registered, hospitalisations not causally related to COVID-19 were excluded.

**Results:** In period 1, there were an estimated 98,170 (95% confidence interval (CI) 96,123-99,928) averted hospitalisations, of which 90,753 (95% CI 88,790-92,531) in period 2, representing 57.0% and 67.9% of all hospital admissions. Estimated averted hospitalisations were lowest for 12-49-year-olds and highest for 70-79-year-olds. More admissions were averted in the Delta period (72.3%) than in the Omicron period (63.4%).

**Conclusion:** COVID-19 vaccination prevented a large number of hospitalisations. Although the estimated number of hospitalisations during the study period could not have realistically occurred due to capacity limits on health care, these findings underline the public health importance of the vaccination campaign to policy makers and the public.

## Introduction

The COVID-19 pandemic has had an enormous negative impact on health and wellbeing worldwide. COVID-19 has directly affected public health with over 600 million confirmed cases to date of which more than 6.5 million deaths (1). In the Netherlands, approximately 8.5 million COVID-19 cases have been confirmed up to August 30, 2022, of which approximately 113,000 have resulted in hospitalisation and more than 22,000 in death (2). Several times during the pandemic, the intensive care units (ICU) were overwhelmed. This imposed pressure on hospital staff, causing a shortage of staff and delaying surgical procedures (3). Also, this pressure increased the need to introduce drastic infection control measures in the community. Although they were effective in controlling the pandemic by decreasing transmission, these control measures also had a negative impact on other aspects of society; for instance, psychological problems in children and adolescents due to social isolation (4) (5), increased prevalence of mental health indicators such as loneliness, anxiety, depression and stress (6) (7), and increased numbers of people facing economic difficulties.

Large-scale COVID-19 vaccination has positively affected public health and reduced the need for restrictions. COVID-19 vaccination has been found effective in limiting infection (8, 9), the transmission of infection (10, 11), the number of hospitalisations and intensive care unit (ICU) admissions (12), and deaths (13, 14). In the Netherlands, the vaccination campaign against COVID-19 started on January 6, 2021 (15). Since then, over 36 million vaccine doses have been administered. A vaccination planning strategy was determined aiming to reduce severe disease. In general, older age groups were eligible before younger age groups, with some exceptions, such as healthcare workers, residents of nursing homes, people living in an institution, and individuals with specific underlying comorbidities (16). All children aged 12-17 could get vaccinated from the beginning of July 2021 (17).

As of today, five types of vaccines have been administered in the Netherlands, Pfizer-BioNTech (COM), Spikevax (MOD), Vaxzevria (VAX), Jcovden (JANSS) and from March 2022, Novavax (NVXD). Two doses are needed to complete the primary vaccination schedule or one dose with a prior SARS-CoV-2 infection, except for JANSS, where one dose is sufficient. From November 18, 2021 (18), booster doses of COM or MOD and from March 25, 2022 (19) booster doses of JANSS have been administered starting with the oldest age groups. A second booster dose of COM or MOD was offered to people over 70 years and some vulnerable health groups from the week of February 28, 2022 (20) and for 60-year-olds from March 26, 2022 (21).

Previous research estimated the substantial beneficial impact of vaccination on health outcomes. An estimated 14.4 million deaths (79%) were prevented in the first year of COVID-19 vaccination based on official reported deaths in 185 countries and territories, or 19.8 million deaths (63%) when estimates were based on excess deaths (22). The percentage of expected deaths in people aged 60 years and older that were averted by vaccination in 33 European countries between December 2020 and November 2021 ranged per country from 6% to 93% with in total for all 33 countries a median of 469,186 deaths (23). A similar study in Italy estimated 79,152 averted hospitalisations (32%) during the roll-out of the vaccination campaign between January and September 2021 (24). Another study showed that in New York City an estimated 8,508 (48.6%) deaths and 48,076 (52.9%) hospitalisations were averted between December 14, 2020 and July 15, 2021 (25).

Quantifying the impact of the vaccination campaign against COVID-19 on hospitalisations helps policy makers and the public to assess the importance of vaccination. With this study we aimed to estimate the number of COVID-19 related hospitalisations averted by the COVID-19 vaccination campaign in the Netherlands.

## Data & Methods

### Data

We estimated the number of COVID-19 related hospitalisations averted by the COVID-19 vaccination campaign from January 6, 2021 through August 30, 2022. We analysed this study period together with a second study period in which all adults had the opportunity to complete their primary series and children aged 12 to 17 were eligible for vaccination, from August 2, 2021 through August 30, 2022. This allowed the impact of the whole vaccination campaign to be differentiated from the impact when all adults had the opportunity to complete the primary series. We present estimates per age group to evaluate the variation in impact of the vaccination campaign.

Vaccine coverage (VC) was determined by registrations in the Dutch COVID-vaccine Information and Monitoring System (CIMS), supplemented with anonymized registrations by the Municipal Health Services (MHS) for those individuals who did not give consent to be registered in CIMS. A completed primary series includes one dose of JANSS, two doses of COM, MOD, NVXD or VAX, or one dose (other than JANSS) following a previous SARS-CoV-2 infection within three months before the dose was administered. Population data per age group is taken from the Dutch population register. Age was calculated as *2021 – birth year*. Individuals with missing birth year were excluded (<0.01%).

All confirmed COVID-19 hospital registrations by the foundation National Intensive Care Evaluation (NICE) were used. All hospitalised persons with a positive SARS-CoV-2 test or CT-confirmed COVID-19 are included in this database. With the rise of Omicron, clinicians increasingly encountered positive SARS-CoV-2 tests in patients hospitalised for reasons that were not directly or indirectly related to COVID-19. Therefore, from January 25, 2022, NICE distinguishes the indication for admission, with four possible indications:

1. Because of COVID-19; COVID-19 is the indication for admission and the patient is treated for it.
2. A combination with COVID-19; COVID-19 is one of the indications for admission; without COVID-19 the patient would not have been hospitalised.
3. Different indication than COVID-19; the patient has COVID-19 but is hospitalized for an unrelated indication.
4. Unknown indication for admission.

Hospital admissions for an indication other than COVID-19 (i.e., indication 3) were excluded from the data. Before January 25, 2022, all hospitalisations were included, since the indication was not recorded. For hospitalisations, age at date of disease onset was estimated as:

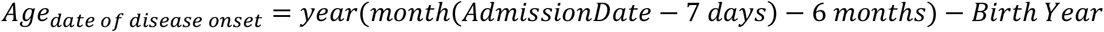

Here, *AdmissionDate* is the hospital admission date and the date of disease onset is estimated by extracting seven days from the admission date, based on the assumption that the median time between date of disease onset and hospitalisation is one week. Because only birth year is known, we extract six months from the date of disease onset, given that approximately half of the persons have aged a year by the middle of the year. We then subtract the birth year to get the estimate of the age at disease onset.

Linked data from CIMS and NICE were used to estimate calendar-time specific vaccine effectiveness (VE) estimates. These estimates with 95% confidence interval (CI) bounds were based on the incidence rate ratio, modelled using a negative binomial regression model, including a natural cubic spline for calendar date and adjusting for birth year in 5-year bands (12). Potential waning and differences in VE between SARS-CoV-2 variants is implicitly included in these VE estimates since the differences in hospitalisation risk associated with vaccination status are modelled over calendar time by including an interaction spline for calendar time and vaccination status. We used the same lag time of seven days to calculate the date of disease onset. To determine age at the date of disease onset for VE estimates, we used the same formula as for hospitalisations that is described above. Hospital admissions for an indication other than COVID-19 were also excluded from the data for estimating the VE.

### Methods

All analyses were done in R (26). We used a study design similar to (23) and (27), with some modifications to include primary series (p), first and second boosters (b1 and b2) vaccine coverage. In short, the method expresses the ratio of averted to observed hospitalisations in terms of VE and VC. We multiply this ratio with the number of observed hospitalisations to get an estimate of the averted hospital admissions. Specifically, we used the following formulas to estimate the number of averted hospitalisations for each vaccination series and per age group:

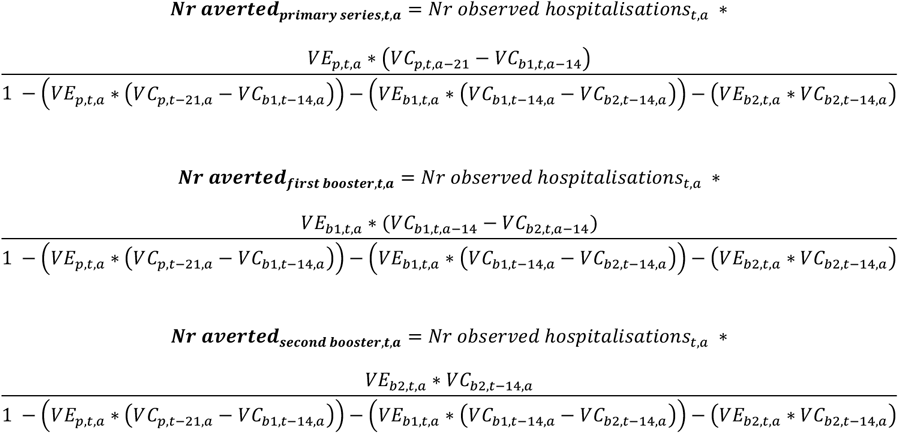

Here, *VE* stands for the vaccine effectiveness against hospitalisation, *VC* for vaccine coverage, *t-x* for a time delay of *x* days and *a* for age group (12-49, 50-59, 60-69, 70-79 and 80+). The delay accounts for the time to immune response after vaccination and median time to hospitalisation after infection. Individuals aged under 12 and individuals whose age is unknown are excluded. We assume a full immune response after 2 weeks for the primary series and after 1 week for the first and second booster, and a time of 1 week between disease onset and disease onset, based on the median delay of reported date of disease onset to hospital admission of confirmed COVID-19 cases, as registered by the Dutch Municipal Health Services. Monte Carlo simulations are used to determine the 95% CI bounds to deal with the uncertainty of VE estimates. The daily VE estimates per age group are used for all point estimates.

The number of averted hospitalisations was estimated per age group from the start of the vaccination campaign in the Netherlands (January 6, 2021) to August 30, 2022 (period 1). The same analysis was carried out for the period between August 2, 2021 and August 30, 2022 (period 2). In period 2, all adults in the Netherlands were eligible for COVID-19 vaccination and had the opportunity to complete the primary series. For this period, observed hospitalisations were used from August 2, 2021 onwards, meaning that the first averted hospitalisations were estimated three weeks later, starting from August 23, 2021, due to the vaccine coverage delay present in the formulas.

We estimated the absolute and relative number of averted hospitalisations per period in which a virus variant was dominant. We define the start of a dominant period as the moment that a variant comprises at least 50% of all test samples of infected individuals that are sequenced by the pathogen surveillance in the Netherlands (28). The periods we use, based on estimated date of disease onset (date of hospitalisation – 7 days), are:

- Wildtype: From the start until February 14, 2021
- Alpha: February 15 until June 27, 2021
- Delta: June 28 until December 26, 2021
- Omicron: December 27, 2021 until August 30, 2022

## Results

Vaccine coverage of the primary series on August 30, 2022, ranges from 73% for age group 12-49 to 94% for 70-79-year-olds (Figure 1.A). At the start of the period 2 on August 2, 2021, when all adults in the Netherlands had the opportunity to complete their primary series, coverage varied from 41% (age group 12-49) to 92% (age group 70-79). In general, the coverage in older age groups was higher compared to younger age groups for primary, first and second booster vaccination (Figure 1.A-C).

**Figure 1.**
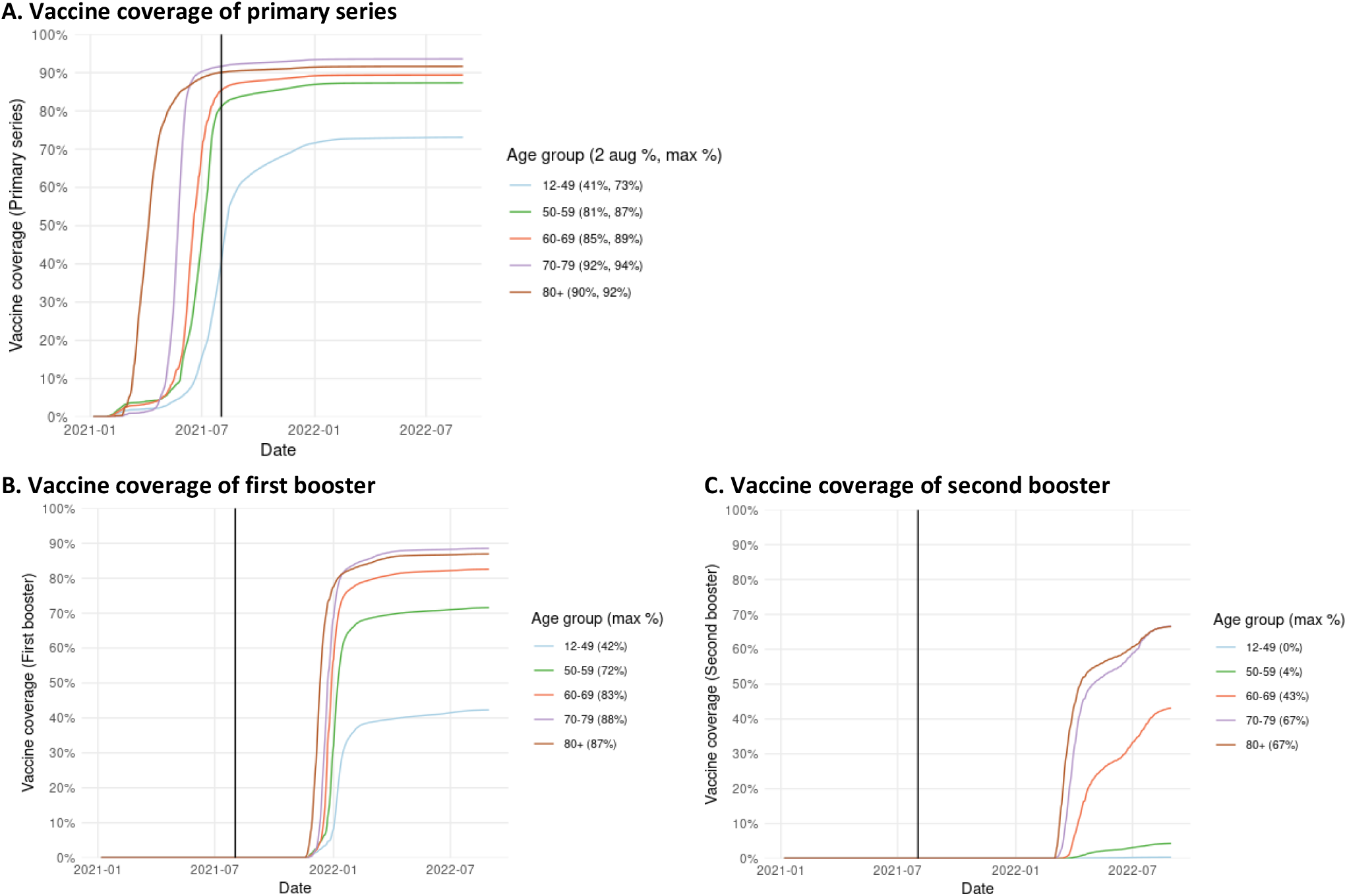
Daily vaccine coverage per age group. The vertical line marks the start of period 2 (August 2, 2021).

In total, 74,074 hospitalisations of individuals of 12 years and older were included in period 1 and 42,930 in period 2. Figure 2 illustrates the weekly number of observed and estimated averted hospitalisations for individuals aged 12 years and older. From the start of the vaccination campaign, an estimated 98,170 hospitalisations (95%CI 96,123 – 99,928) have been averted, with most of them in period 2 (90,753; 95%CI 88,790 – 92,531). These numbers equal 57.0% (56.5%-57.4%) and 67.9% (67.4%-68.3%) of all hospital admissions for respectively the first and second study period.

**Figure 2.**
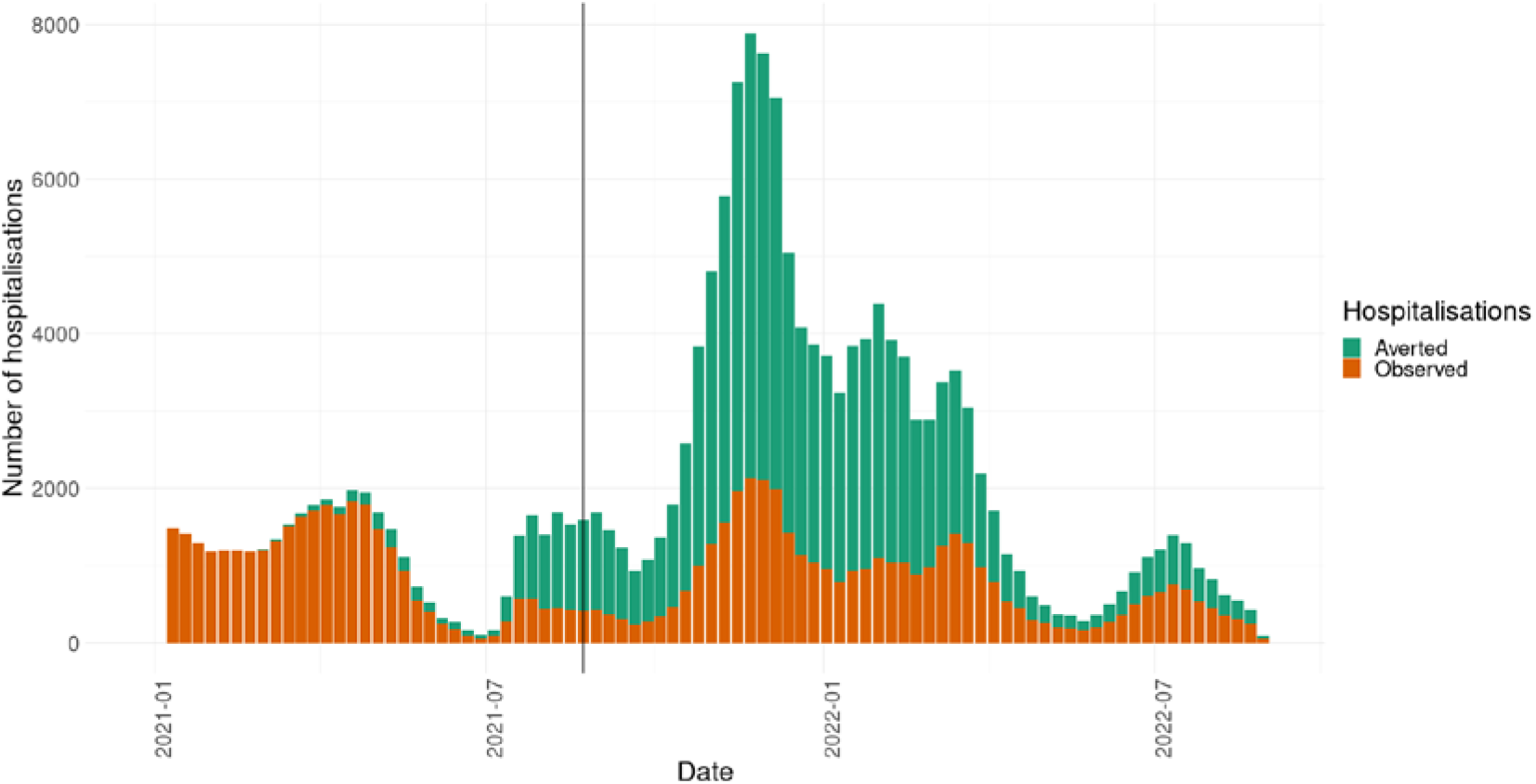
Total weekly number of observed and estimated averted hospitalisations for individuals aged 12 years and older. The vertical line marks the cut-off for estimated averted hospitalisations in period 2 (August 23, 2021, 3 weeks after the start of the study period, accounting for the delay in the time to immune response following completion of the primary vaccination series and the median time to hospitalisation following infection).

Figure 3 summarises the observed and estimated averted hospitalisations per age group for the first study period from January 6, 2021, and for the second study period from August 2, 2021. The number of estimated averted hospitalisations was highest for age group 70-79 years, both absolute (32,483, of which 30,268 in the second study period) and relative to observed hospitalisations (63.5% from the start and 73.3% in the second study period). The lowest numbers were observed for age group 12-49 years: 7,607 (37.3%) estimated averted hospitalisations, of which 7,255 (49.9%) in the second study period.

**Figure 3.**
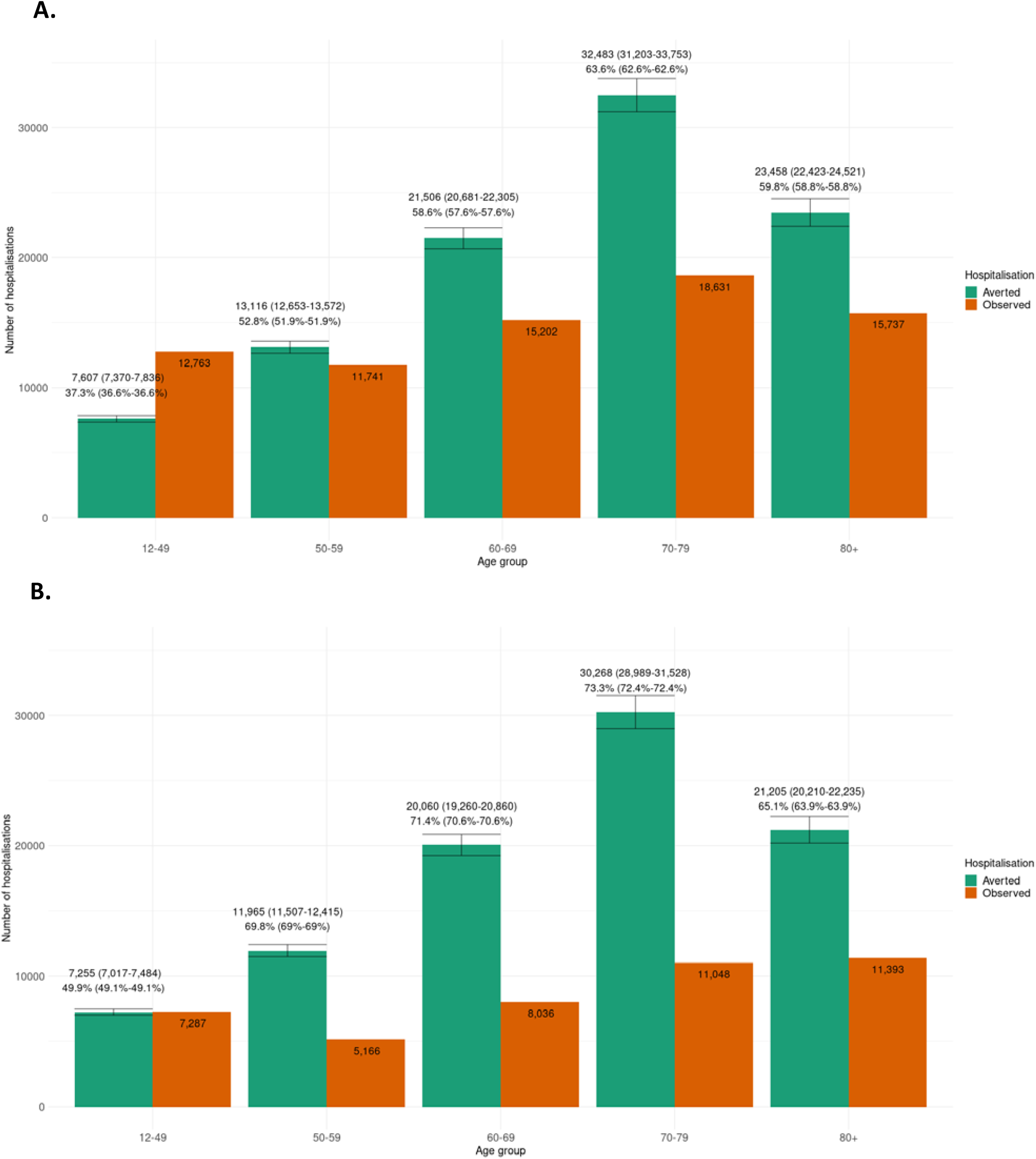
Percentage and absolute number of estimated averted hospitalisations with 95% CI bounds, and the number of observed hospitalisations, per age group. A. Period 1 (January 6, 2021 until August 30, 2022) B. Period 2 (August 2, 2021 until August 30, 2022)

During the Alpha period, an estimated 1,826 (8.1%; 95%CI 1,767 – 1,876; 7.8% - 8.3%) hospitalisations were averted by vaccination (Figure 4). The largest number of hospitalisations was averted during the Delta period, both in absolute (57,395; 95%CI 55,918 – 58,723) as well as relative (72.3%, 71.8% - 72.8%) terms. In the Omicron period, an estimated 38,949 (63.4%) admissions were averted (95%CI 37,749 – 40,152, 62.7% - 64.1%).

**Figure 4.**
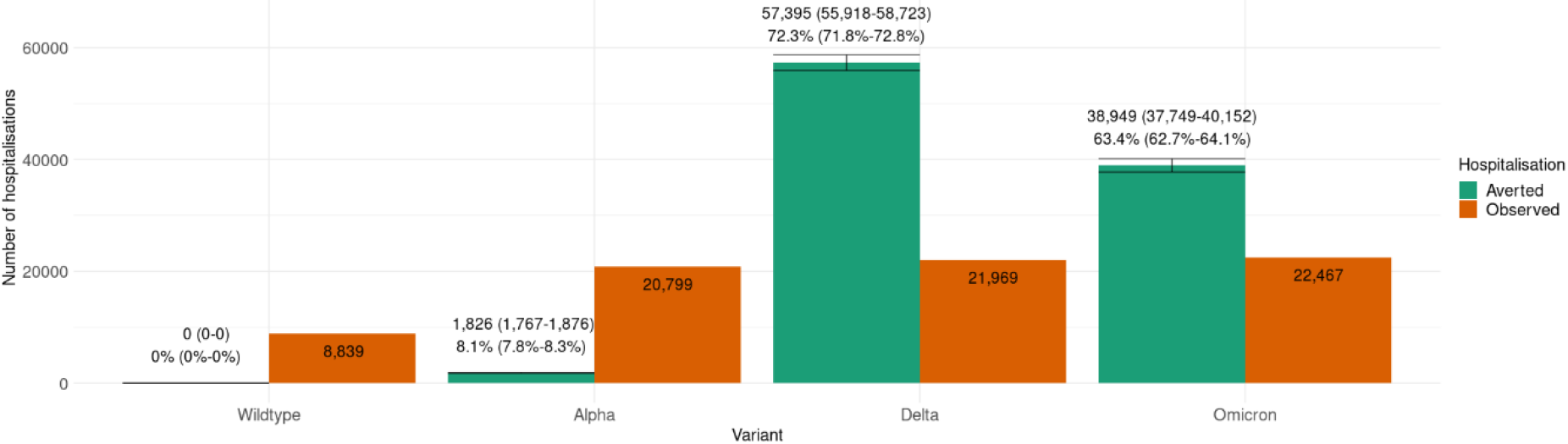
Number and percentage of estimated averted hospitalisations and number of observed hospitalisations per variant, from the start of the vaccination campaign (January 6, 2021) until August 30, 2022

## Discussion

We aimed to quantify the impact of COVID-19 vaccination in the Netherlands in terms of averted hospitalisations. Results show that the vaccination campaign has had a considerable, positive impact on an important indicator of COVID-19 health burden – admission to hospital – which reinforces the investment in the vaccination programme for preventing serious disease at the patient level, reducing the associated burden on the healthcare system, and retaining capacity to admit non-COVID-19 patients. An estimated 98,170 (57.0%) admissions have been averted from the start of the vaccination campaign, of which 90,753 (67.9%) in the period where all adults had the opportunity to complete the primary series and children aged 12 to 17 were eligible for vaccination (period 2).

An upward trend can be seen in the fraction of averted hospitalisations from younger to older age groups, with a peak for age group 70-79. This age trend is similar to the vaccine coverage, which is higher for older age groups, and highest for age group 70-79. The greatest percentage of hospital admissions were averted in the Delta period. VE decreased significantly with the emergence of Omicron. While VE against hospitalisation remained much higher than against infection, a drop in VE was still observed around the start of 2022.

The difference in the percentage averted admissions between the period from the start of the vaccination campaign (period 1) and the period in which all adults in the Netherlands had the opportunity to complete the primary series (period 2) is substantial, with a total difference of over 10%. This underlines the need for a rapid roll-out of the vaccination campaign, especially in times when the number of observed hospital admissions is high. The very large difference in the relative number of averted hospitalisations between the Alpha period (8.1%) and the Delta and Omicron periods (72.3% and 63.4%), with the Alpha period comprising a large part of initial roll-out, highlights the importance of a rapid roll-out.

Comparing our relative estimates to relative estimates of averted hospitalisations or deaths in previous research is not straightforward, because the calculations depend on several factors. Firstly, outcomes are affected by the VE estimates used. The VE against death is higher than the VE against hospitalisation, which leads to a lower expected percentage averted hospitalisations compared to averted deaths. Secondly, a higher vaccine coverage leads to a higher estimated percentage of averted admissions. In the Netherlands, coverage is relatively high, thus we would expect a higher estimate. Lastly, the number of observed hospitalisations, the timing of peaks in observed hospitalisations, and the length of the study period affect the relative estimates. Nevertheless, our estimate in period 1 (57.0%) is similar to the estimated global reduction in deaths of 63% in (22), the estimate of 50% averted deaths for neighbouring country Belgium (which had similar vaccine coverage) (23), and the estimate of 52.9% averted hospitalisations for New York (25). It is much higher than the estimated 32% averted hospitalisations for Italy (24), which can be explained by differences in study period; in this study, the study period consisted of mainly the roll-out period of the vaccination campaign (January 2021 to September 2021).

The current study has several limitations. The CIMS data, from which calendar-time specific VE estimates were derived, did not contain vaccination records for individuals who did not consent to share their data (in total around 6.2% of all administered doses). This will have resulted in misclassification of some vaccinated individuals as unvaccinated, leading to an underestimation of VE estimates (12). Secondly, the VE estimates were not adjusted for comorbidities, as this information could not be individually linked to the hospital and vaccination data. Presumably, this has also resulted in conservative VE estimates, assuming that individuals with a higher risk of severe illness from COVID-19 were invited for vaccination earlier and were more likely to accept vaccination.

Similarly, because previous infections could not be individually linked to the hospital and vaccination data, it was not possible to adjust for previous infections in the VE analyses. Over time, people regardless of vaccination status accrued infection-induced or hybrid immunity. This phenomenon may have led to underestimation of VE if unvaccinated individuals reached infection-induced immunity earlier and more frequently (29). Additionally, we do not consider indirect effects of vaccination in our analysis. Vaccination not only protects against severe COVID-19, but also reduces the probability of infecting other individuals (11, 30). This leads to fewer hospitalisations, and therefore not taking this into account makes the estimate more conservative.

Additional limitations may have resulted in an overestimation of averted hospitalisations. Firstly, we did not remove once-hospitalised persons (either observed or averted) from the population at risk for future hospitalisations, thus allowing for individuals to be admitted to the hospital more than once. In practice, the probability of being hospitalised after a prior hospital admission is likely lower than without a prior hospital admission. Secondly, the indication for COVID-19 hospitalisation was unknown until January 25, 2022. Hospitalisations with a positive SARS-CoV-2 test – but not due to COVID-19 – could thus not be excluded before this date. Because clinicians increasingly saw patients being hospitalised with a positive SARS-CoV-2 test, but whose indication for admission was not because of COVID-19, once the Omicron variant became dominant, registration of the indication of admission was added not long afterwards. Therefore, we assumed the impact on our estimates was limited. In addition, for the VE estimates the indication was unknown until January 25, 2022. Under the assumption that vaccination has no effect on the incidence of hospitalisations with – but not due to COVID-19 – VE would be underestimated. We therefore expect that this limitation has more impact on the relative number of estimated averted hospitalisations, leading to underestimation, than on the absolute number, possibly leading to minor overestimation. The same applies to hospital admissions after January 25, 2022 with an unknown indication for admission.

Determining the impact of the COVID-19 vaccination campaign is challenging, since the counterfactual is unknown; we do not know what would have happened in the absence of the vaccination campaign. In practice, hospitals would not have been capable of admitting the enormous estimated number of patients needing hospital care, since their capacity would then have been exceeded. It is likely that stricter control measures would have been required to limit the total number of COVID-19 hospitalisations and/or spread admissions out over a longer time period. In our study, however, we estimated the impact under the assumption that public health measures and compliance to these measures would have been identical to the actual measures that had been imposed, and we assumed that there was no limit in hospital bed and staff capacity. Nevertheless, quantification of the effect of the vaccination campaign in terms of averted hospitalisations demonstrates its considerable positive impact on public health.

## Conclusion

From January 6, 2021 until August 30, 2022 an estimated 98,170 (95% CI 96,123-99,928) COVID-19 hospitalizations were averted by the SARS-CoV-2 vaccination campaigns, relative to 74,074 observed hospitalizations. The relative and absolute number of averted hospitalizations was most pronounced for age group 70-79 and during the Delta period. COVID-19 vaccination prevented a considerable burden of morbidity by reducing the number of COVID-19 hospitalisations. By doing so, it helped to improve access to healthcare for both COVID-19 patients as well as non-COVID-19 patients, due to less pressure on hospitals and healthcare workers.

## Data Availability

The primary data is not publicly available. Aggregated tables of these data are made available weekly at the website of the RIVM.

## Members of the RIVM COVID-19 surveillance and epidemiology team

Albert Jan van Hoek, Agnetha Hofhuis, Amber Maxwell, Annabel Niessen, Anne Teirlinck, Anne-Wil Valk, Carolien Verstraten, Claudia Laarman, Danytza Berry, Daphne van Wees, Dimphey van Meijeren, Don Klinkenberg, Eric Vos, Femke Jongenotter, Fleur Petit, Frederika Dijkstra, Guido Willekens, Irene Veldhuijzen, Jacco Wallinga, Jan van de Kassteele, Janneke van Heereveld, Janneke Heijne, Jantien Backer, Jeanet Kemmeren, Kirsten Bulsink, Kylie Ainslie, Liselotte van Asten, Liz Jenniskens, Lieke Wielders, Loes Soetens, Maarten Mulder, Maarten Schipper, Manon Haverkate, Marit de Lange, Marit Middeldorp, Naomi Smorenburg, Nienke Neppelenbroek, Patrick van den Berg, Pieter de Boer, Priscila de Oliveira Bressane Lima, Rianne van Gageldonk-Lafeber, Rolina van Gaalen, Steven Nijman, Stijn Andeweg, Susan Lanooij, Susan van den Hof, Tara Smit, Thomas Dalhuisen, Tjarda Boere

## References

1. World Health Organization. WHO Coronavirus (COVID-19) Dashboard: World Health Organization; 2022 [Available from: https://covid19.who.int/.

2. Dong E, Du H, Gardner L. An interactive web-based dashboard to track COVID-19 in real time. The Lancet Infectious Diseases. 2020;20(5):533–4.

3. de Wit GA, Oosterhoff M, Kouwenberg LHJA, Rotteveel AH, van Vliet ED, Janssen K, et al. De gezondheidsgevolgen van uitgestelde operaties tijdens de corona-pandemie. Schattingen voor 2020 en 2021. National Institute for Public Health and the Environment; 2022.

4. Racine N, McArthur BA, Cooke JE, Eirich R, Zhu J, Madigan S. Global Prevalence of Depressive and Anxiety Symptoms in Children and Adolescents During COVID-19: A Meta-analysis. JAMA Pediatrics. 2021;175(11):1142–50.

5. Luijten MAJ, van Muilekom MM, Teela L, Polderman TJC, Terwee CB, Zijlmans J, et al. The impact of lockdown during the COVID-19 pandemic on mental and social health of children and adolescents. Quality of Life Research. 2021;30(10):2795–804.

6. Daly M, Robinson E. Psychological distress and adaptation to the COVID-19 crisis in the United States. Journal of Psychiatric Research. 2021;136:603–9.

7. van Tilburg TG, Steinmetz S, Stolte E, van der Roest H, de Vries DH. Loneliness and Mental Health During the COVID-19 Pandemic: A Study Among Dutch Older Adults. The Journals of Gerontology: Series B. 2020;76(7):e249–e55.

8. Lopez Bernal J, Andrews N, Gower C, Gallagher E, Simmons R, Thelwall S, et al. Effectiveness of Covid-19 Vaccines against the B.1.617.2 (Delta) Variant. N Engl J Med. 2021;385(7):585–94.

9. Andeweg SP, de Gier B, Eggink D, van den Ende C, van Maarseveen N, Ali L, et al. Protection of COVID-19 vaccination and previous infection against Omicron BA.1, BA.2 and Delta SARS-CoV-2 infections. Nature Communications. 2022;13(1):4738.

10. de Gier B, Andeweg S, Joosten R, ter Schegget R, Smorenburg N, van de Kassteele J, et al. Vaccine effectiveness against SARS-CoV-2 transmission and infections among household and other close contacts of confirmed cases, the Netherlands, February to May 2021. Eurosurveillance. 2021;26(31):2100640.

11. de Gier B, Andeweg S, Backer JA, RIVM COVID-19 surveillance epidemiology team, Hahné SJM, van den Hof S, et al. Vaccine effectiveness against SARS-CoV-2 transmission to household contacts during dominance of Delta variant (B.1.617.2), the Netherlands, August to September 2021. Eurosurveillance. 2021;26(44):2100977.

12. de Gier B, Kooijman M, Kemmeren J, de Keizer N, Dongelmans D, van Iersel SCJL, et al. COVID-19 vaccine effectiveness against hospitalizations and ICU admissions in the Netherlands, April-August 2021. medRxiv. 2021:2021.09.15.21263613.

13. Sheikh A, Robertson C, Taylor B. BNT162b2 and ChAdOx1 nCoV-19 Vaccine Effectiveness against Death from the Delta Variant. New England Journal of Medicine. 2021;385(23):2195–7.

14. de Gier B, van Asten L, Boere T, van Werkhoven H, van Roon A, van den Ende C, et al. COVID-19 vaccine effectiveness against mortality and risk of death from other causes after COVID-19 vaccination, the Netherlands, January 2021-January 2022. medRxiv. 2022:2022.07.21.22277831.

15. Valk A, van Meijeren DL, Smorenburg N, Neppelenbroek NJM, van Iersel SCJL, de Bruijn S, et al. Vaccinatiegraad COVID-19 vaccinatie Nederland, 2021. National Institute for Public Health and the Environment; 2022.

16. Pluijmaekers AJM, de Melker HE. The National Immunisation Programme in the Netherlands. Surveillance and developments in 2021-2022. National Institute for Public Health and the Environment; 2022.

17. Jongeren van 12 tot en met 17 jaar kunnen zich vanaf begin juli laten vaccineren 2021 [Available from: https://www.rijksoverheid.nl/actueel/nieuws/2021/06/30/jongeren-van-12-tot-en-met-17-jaar-kunnen-zich-vanaf-begin-juli-laten-vaccineren.

18. Boosterprik versneld van start 2021 [Available from: https://www.rijksoverheid.nl/actueel/nieuws/2021/11/12/boosterprik-voor-60-plussers-versneld-van-start.

19. Janssen booster possible as of 25 March 2022 [Available from: https://www.rivm.nl/en/news/janssen-booster-possible-as-of-25-march.

20. Extra coronaprik voor 70-plussers, bewoners van verpleeghuizen en mensen met een ernstig verminderde weerstand 2022 [Available from: https://www.rijksoverheid.nl/actueel/nieuws/2022/02/24/extra-coronaprik-voor-70-plussers-bewoners-van-verpleeghuizen-en-mensen-met-een-ernstig-verminderde-weerstand.

21. Extra coronaprik nu ook voor 60-plussers 2022 [Available from: https://www.rijksoverheid.nl/actueel/nieuws/2022/03/25/extra-coronaprik-nu-ook-voor-60-plussers.

22. Watson OJ, Barnsley G, Toor J, Hogan AB, Winskill P, Ghani AC. Global impact of the first year of COVID-19 vaccination: a mathematical modelling study. The Lancet Infectious Diseases. 2022;22(9):1293–302.

23. Meslé MM, Brown J, Mook P, Hagan J, Pastore R, Bundle N, et al. Estimated number of deaths directly averted in people 60 years and older as a result of COVID-19 vaccination in the WHO European Region, December 2020 to November 2021. Eurosurveillance. 2021;26(47):2101021.

24. Sacco C, Mateo-Urdiales A, Petrone D, Spuri M, Fabiani M, Vescio MF, et al. Estimating averted COVID-19 cases, hospitalisations, intensive care unit admissions and deaths by COVID-19 vaccination, Italy, January−September 2021. Eurosurveillance. 2021;26(47):2101001.

25. Shoukat A, Vilches TN, Moghadas SM, Sah P, Schneider EC, Shaff J, et al. Lives saved and hospitalizations averted by COVID-19 vaccination in New York City: a modeling study. The Lancet Regional Health - Americas. 2022;5:100085.

26. R Core Team. R: A language and environment for statistical computing. Vienna, Austria: R Foundation for Statistical Computing; 2022.

27. Machado A, Mazagatos C, Dijkstra F, Kislaya I, Gherasim A, McDonald SA, et al. Impact of influenza vaccination programmes among the elderly population on primary care, Portugal, Spain and the Netherlands: 2015/16 to 2017/18 influenza seasons. Eurosurveillance. 2019;24(45):1900268.

28. Covid-19 rapportage van SARS-CoV-2 varianten in Nederland via de aselecte steekproef van RT-PCR positieve monsters in de nationale kiemsurveillance. 2022.

29. Kahn R, Schrag SJ, Verani JR, Lipsitch M. Identifying and Alleviating Bias Due to Differential Depletion of Susceptible People in Postmarketing Evaluations of COVID-19 Vaccines. American Journal of Epidemiology. 2022;191(5):800–11.

30. Lyngse FP, Mortensen LH, Denwood MJ, Christiansen LE, Møller CH, Skov RL, et al. Household transmission of the SARS-CoV-2 Omicron variant in Denmark. Nature Communications. 2022;13(1):5573.

